# Reproducing the long term predictions from Imperial College CovidSim Report 9

**DOI:** 10.1101/2020.06.18.20135004

**Authors:** Ken Rice, Ben Wynne, Victoria Martin, Graeme Ackland

## Abstract

We present calculations using the CovidSim code which implements the Imperial College individual-based model of the COVID epidemic. Using the parameterization assumed in March 2020, we reproduce the predictions presented to inform UK government policy in March 2020. We find that CovidSim would have given a good forecast of the subsequent data if a higher initial value of R0 had been assumed. We then investigate further the whole trajectory of the epidemic, presenting results not previously published. We find that while prompt interventions are highly effective at reducing peak ICU demand, none of the proposed mitigation strategies reduces the predicted total number of deaths below 200,000. Surprisingly, some interventions such as school closures were predicted to increase the projected total number of deaths.

## 2 Introduction

The UK national response to the coronavirus crisis has been widely reported as being primarily led by modelling based on the work of (5), although other models have been considered_1_. The key paper (4), which we will refer to as “Report 9”, investigated a number of scenarios using this code with the best parameterisation available at the time. Contrary to popular perception, the full lockdown which was then implemented was not specifically modelled in this work. As the pandemic has progressed, the parameterisation has been continually improved with new data as it arrives.

The main conclusions of Report 9 were not especially surprising. COVID has a morbidity around 1% (9), so an epidemic in a susceptible population of 70M people would cause many hundreds of thousands of deaths. In early-March there may have been a case-doubling time of around 3 days in the UK (12), meaning that within a week COVID cases could go from accounting for a minority of available ICU spaces, to exceeding capacity. Furthermore, with an onset delay of over a week, and limited or delayed testing and reporting in place, there would be very little measurable warning of the explosion in ICU demand.

The original code used for Report 9 has not been released, however the Ferguson group has led an effort with Microsoft, github and the Royal Society RAMP-initiative to recreate the model in the open-sourced CovidSim code(8). CovidSim faithfully replicated the original code, and recently the results presented in Report 9 were independently replicated(1).

CovidSim is an individual-based code that includes a detailed geographic network, considers schools, universities, and places of work, and is parameterised using the best available expert clinical and behavioural evidence. The model has the required complexity to consider non-pharmaceutical interventions (NPIs), specifically home isolation of suspect cases (CI), home quarantine of family members (HQ), general social distancing (SD), and social distancing of those over 70 (SDOL70). It further considered “place closures” (PC), specifically the closure of schools and universities. More details of these NPIs are provided in Table 2 of Report 9, which we reproduce in Appendix Figure 5.

Report 9 presented both mitigation and suppression strategies, but we focus here on the mitigation strategies. One counter-intuitive result presented in Report 9 (their Table 3 and Table A1) is the prediction that once all other considered interventions were in place, the additional closure of schools and universities would **increase** the total number of deaths. Similarly, adding general social distancing (SD) to a scenario involving case isolation and household quarantine, was also projected to increase the total number of deaths.

Very recently, the detailed inputs used for Report 9 were released. In this paper, we reproduce the main results from Report 9, and explain why, in the framework of the model, these counter-intuitive results were obtained.

## 3 Methods

The CovidSim model is developed from an influenza pandemic model(5; 6; 10). We used the version recovered from github(8), tagged version 0.14.0 + additional patches dated before 03-06-2020. Input files relevant to Report 9 were supplied by Ferguson et al.(3).

We chose not to attempt to re-parameterise the model. CovidSim contains a huge number of parameters derived from expert judgement, and so it is not well suited to data-driven re-parameterisation using Bayesian inference or related techniques. The epidemiological assumptions underlying our results are identical to those of Ferguson et al., the purpose of this paper is to investigate more fully the consequences of those assumption.

## 4 Results

The result tables for the scenarios presented in the original report were straight-forwardly reproduced by averaging over 10 simulation runs with the same random number seeds as used in Report 9. The simulations are run for 800 days, with day 1 being 01 January 2020. The intervention period lasts for 3 months (91 days), with some interventions extended for an additional 30 days. The mitigation scenarios in Report 9 considered *R*_0_ = 2.2 and *R*_0_ = 2.4, but we initially only considered *R*_0_ = 2.4.

As highlighted in (8) the results we obtain here are not precisely identical to those in Report 9, since they are an average over 10 stochastic realisations, the population dataset has changed to one that is open-source, and the algorithm used to generate the household-to-place network has been modified to be deterministic. The stochasticity gives a variance around 5% in total deaths and ICU demand, which explains the discrepancies with Report 9. More significant is the uncertainty of the timing of the peak of the infections, which is around ± 5 days.

We compare these predictions to the death rates from the actual trajectory of the disease(11). We note that NHS England stopped publishing critical bed occupancy in March 2020(2), so it is not possible to compare ICU data from the model with reality.

To understand why, for example, closing schools and universities increases total deaths within the model, we consider two possibilities.

Firstly, reduced contact at school leads to increased contact at home. Therefore infection is transferred from low-risk children to high-risk adults. We investigated this by examining the effect of changing the ”Relative household contact rate after closure” parameter. In Figure 1 it is clear that variation due to the value of this parameter is insignificant compared to the overall effect of adding school closures^2^ to the other interventions. Therefore we reject this hypothesis.

**Figure 1:**
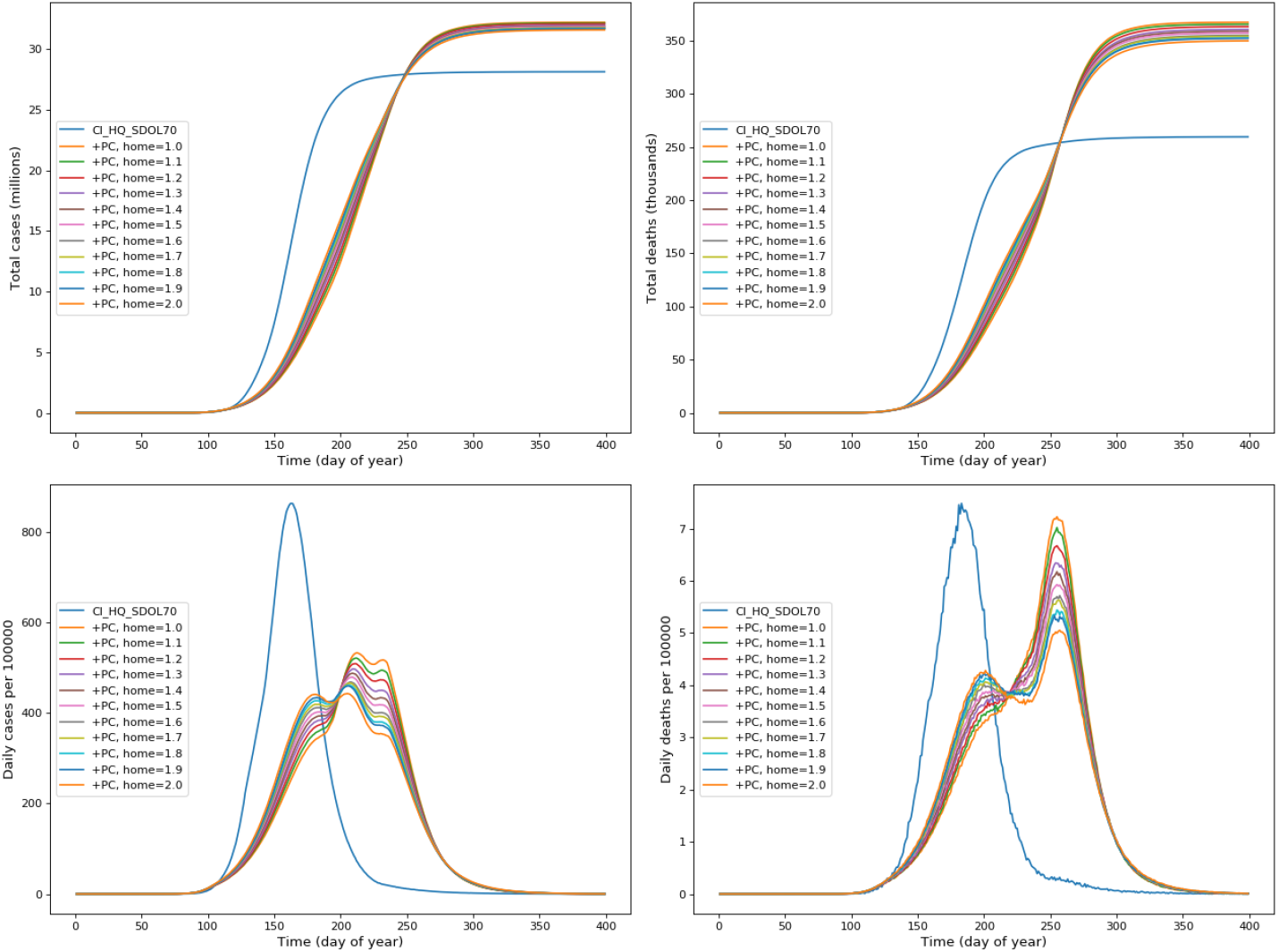
Comparison of simulation results over time, for the CI_HQ_SDOL70 and PC_CI_HQ_SDOL70 intervention scenarios. Interventions are triggered by reaching 100 cumulative ICU cases. In the scenario including Place Closure (PC), the value of the relative household contact parameter is varied from 1.0 to 2.0. Regardless of this variation, the additional PC intervention always leads to an increase in total cases and deaths.

Secondly, place closures suppress the first wave, but when the interventions are lifted, there is still a large population of people who are susceptible and a substantial number of people who are infected. This then leads to a second wave of infections that can result in more deaths, but at a later time.

In Table 1 we show the critical care (ICU) bed demand, while in Table 2 we show total deaths, both using the same mitigation scenarios as presented in Report 9. As in Report 9, for each mitigation scenario we consider a range of ICU triggers. In Table 1 we report the peak ICU bed demand across the full simulation for each trigger, as was presented in Report 9, but also include the peak ICU bed demand during the period of the intervention (labelled **(Int)**). The latter we define as the period during which general social distancing (SD) was in place, when implemented.

**Table 1:**
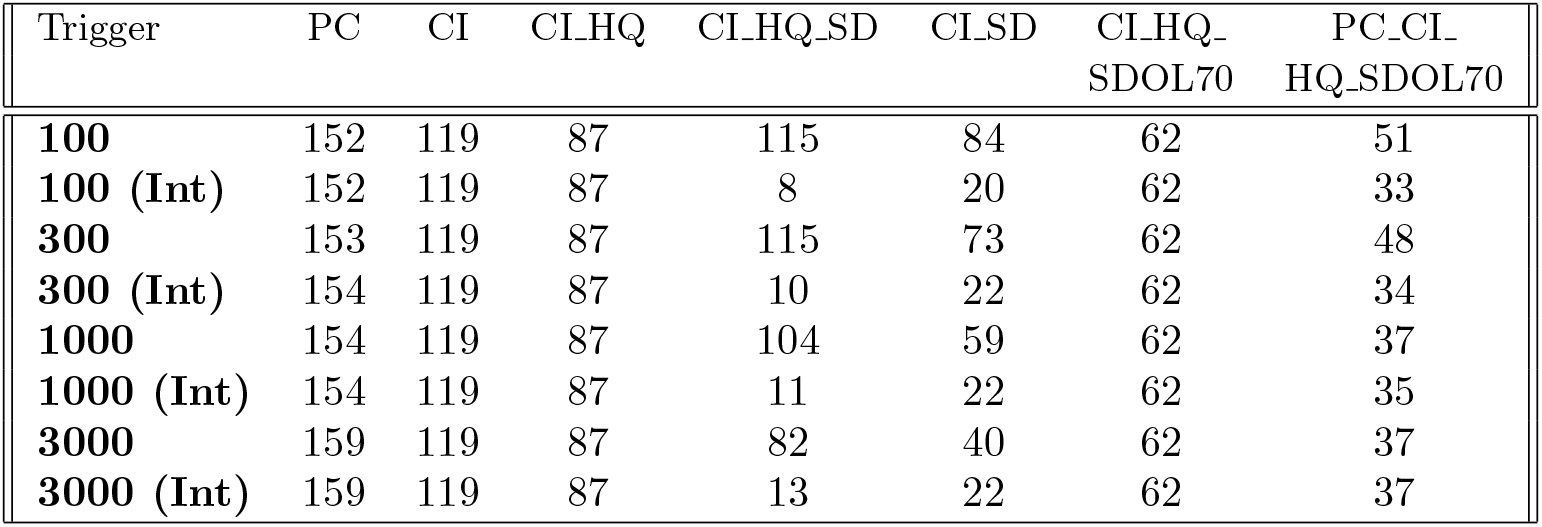
Table showing peak ICU bed demand (UK-wide in thousands) for different intervention scenarios and different ICU triggers. For each trigger, we show the peak ICU bed demand across the full simulation, and during the first period when the interventions were in place (labelled **(Int)**).

**Table 2:**
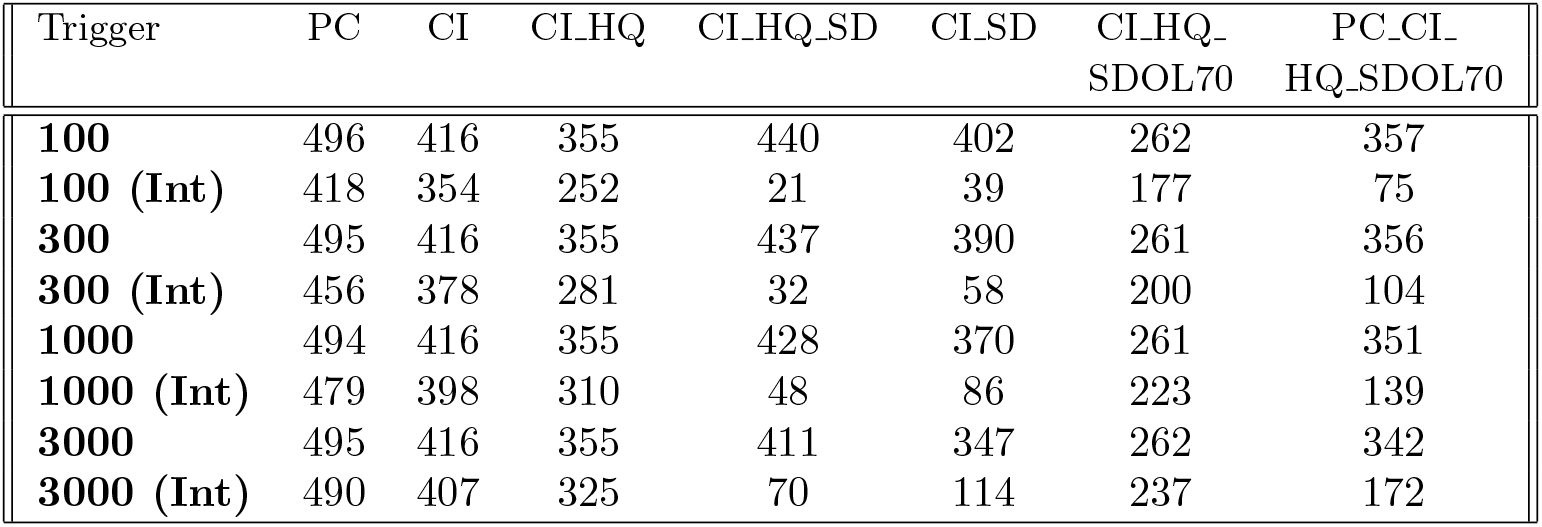
Table showing total deaths (UK-wide, in thousands) for different intervention scenarios and different ICU triggers. For each trigger, we show the total deaths across the full simulation, and also up to the time at which interventions were first lifted (labelled **(Int)**).

In Table 2 we also report the total number of deaths across the entire simulation, and also the number of deaths at the time when the interventions were lifted, again defined as the time at which general social distancing was lifted, if implemented.

The full simulation numbers we present in Tables 1 and 2 are essentially the same as those presented in Table A1 in Report 9. As discussed earlier, the small difference between our numbers and those presented in Report 9 are probably because these are averaged over 10 stochastic realisations, the population dataset is slightly different, and the algorithm for generating the household-to-place network was changed to make it deterministic. Table 2 also illustrates the counter-intuitive result that adding PC to CI_HQ_SDOL70 increases the total number of deaths across the full simulation, as does adding SD to CI_HQ.

What’s clear from Tables 1 and 2 is that in some mitigation scenarios peak ICU demand, and most deaths, occur during the period when the intervention is in place. There are, however, other scenarios where the peak ICU demand, and total deaths, during the period of the intervention is far smaller than outside this period.

The reason for this is illustrated in Figure 2. The solid lines are the same mitigation scenarios as presented in Figure 2 of Report 9. We also show some additional scenarios (dashed lines) not shown in Figure 2 of Report 9, but which are included in Tables 1 and 2 and also in the Tables in Report 9.

**Figure 2:**
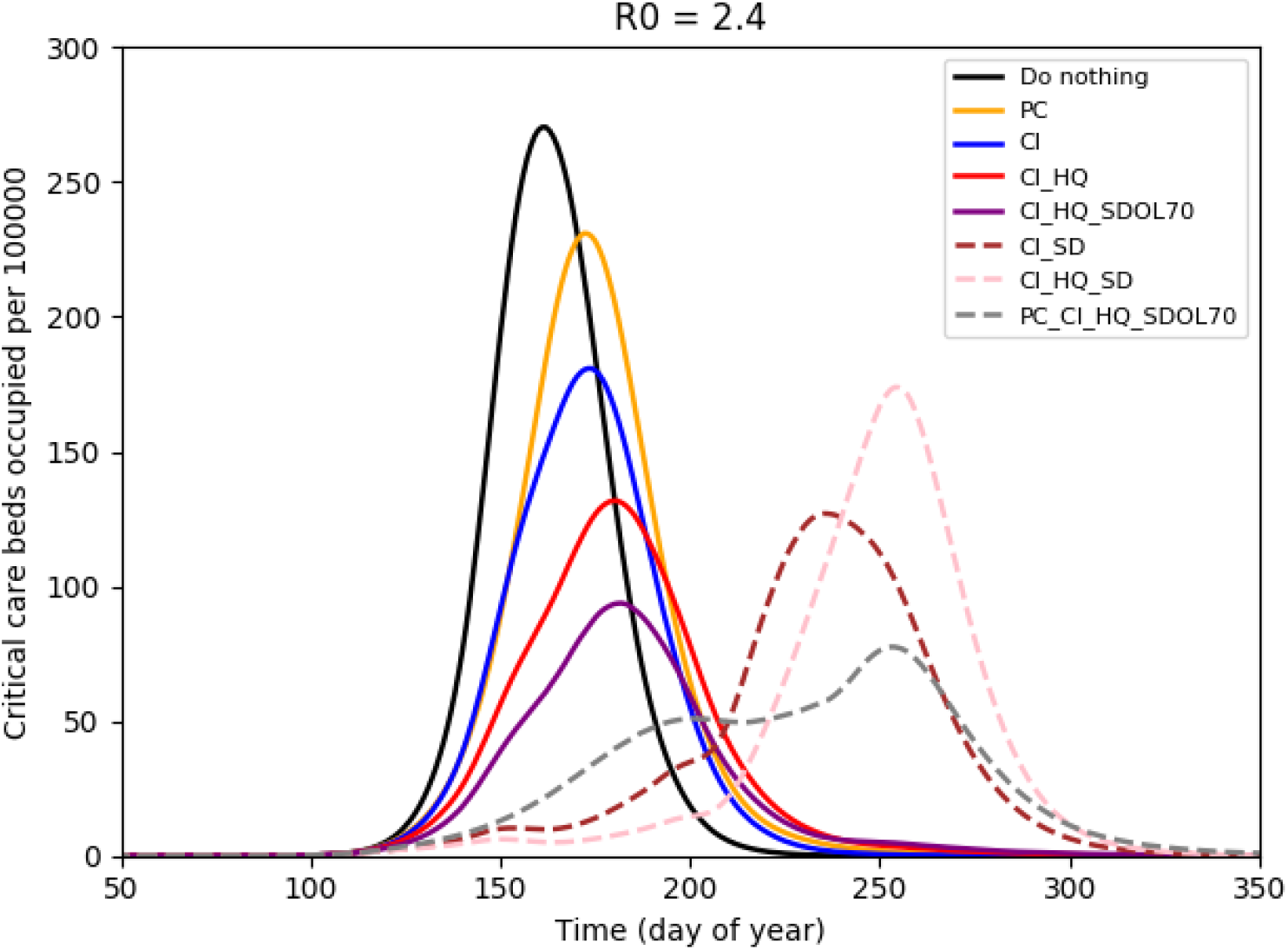
Equivalent of Figure 2 from report 9, for *R*_0_ = 2.4. The solid lines are the same scenarios as presented in Figure 2 of report 9. We also include three additional scenarios (dashed lines) not presented in Figure 2 of report 9, but which are presented in their Tables and in Tables 1 and 2 here. These illustrate why adding place closures (PC) to a scenario with case isolation (CI), household quarantine (HQ) and social distancing of those over 70 (SDOL70) can lead to more deaths than the equivalent scenario without place closures. Doing so suppresses the infection when the interventions are present, but leads to a second wave when they are lifted, which happens on around day 200. The total number of deaths in the CI_HQ_SDOL70 scenario is 260,000, while for PC_CI_HQ_SDOL70 it is 350,000. So the latter has more deaths even though the peak ICU bed demand is lower. A similar effect can be seen by comparing scenarios with general social distancing (SD) with equivalent scenarios without SD. For example, the second wave peak in the CI_HQ_SD scenario is actually higher than the first wave peak in the CI_HQ scenario.

In the simulations presented here, the main interventions are in place for 3 months and end on about day 200 (some interventions are extended for an additional 30 days). Figure 2 shows that some intervention scenarios lead to a single wave that occurs during the period in which the interventions are in place. Hence, the peak ICU bed demand occurs during this period, as do most deaths.

There are, however, some interventions that suppress the infection so that there is then a second wave once the interventions are lifted. For example, adding place closures to case isolation, household quarantine, and social distancing of those over 70 substantially suppresses the infection during the intervention period when compared to the same scenario without place closures. However, this suppression then leads to a second wave with a higher peak ICU bed demand than during the intervention period, and total deaths that exceed that of the same scenario without place closures.

We therefore conclude that the somewhat counter-intuitive results presented in Report 9 are a consequence of the addition of some interventions suppressing the first wave so that a second wave, occurring after the interventions have lifted, then leads to a total number of deaths that exceeds the total for the equivalent scenario without this additional intervention.

A similar effect can be seen in some of the scenarios involving general social distancing (SD). For example, adding general social distancing to case isolation and household quarantine also strongly suppresses the infection during the intervention period, but then leads to a second wave that actually has a higher peak ICU demand than for the equivalent scenario without general social distancing.

Figure 3 provides an explanation for how place closure interventions affect the second wave, and why an extra intervention may result in more deaths than the equivalent scenario without this intervention. In the CI_HQ_SDOL70 scenario, without closures, a single peak of cases is seen. The data is broken-down into age groups, showing that younger people contribute most to the total cases, but that deaths come primarily from older groups. Adding the place closure intervention (and keeping all other things constant) gives the behaviour shown in the second row of plots. The initial peak is greatly suppressed, but the end of closures seems to prompt a second peak of cases amongst younger people. This then leads to a third, more deadly, peak of cases affecting the elderly when SDOL70 is removed. The net effect is that there is a postponement in the number of infections in the younger age groups, which increases the number of infections, and hence deaths, in the older age groups.

**Figure 3:**
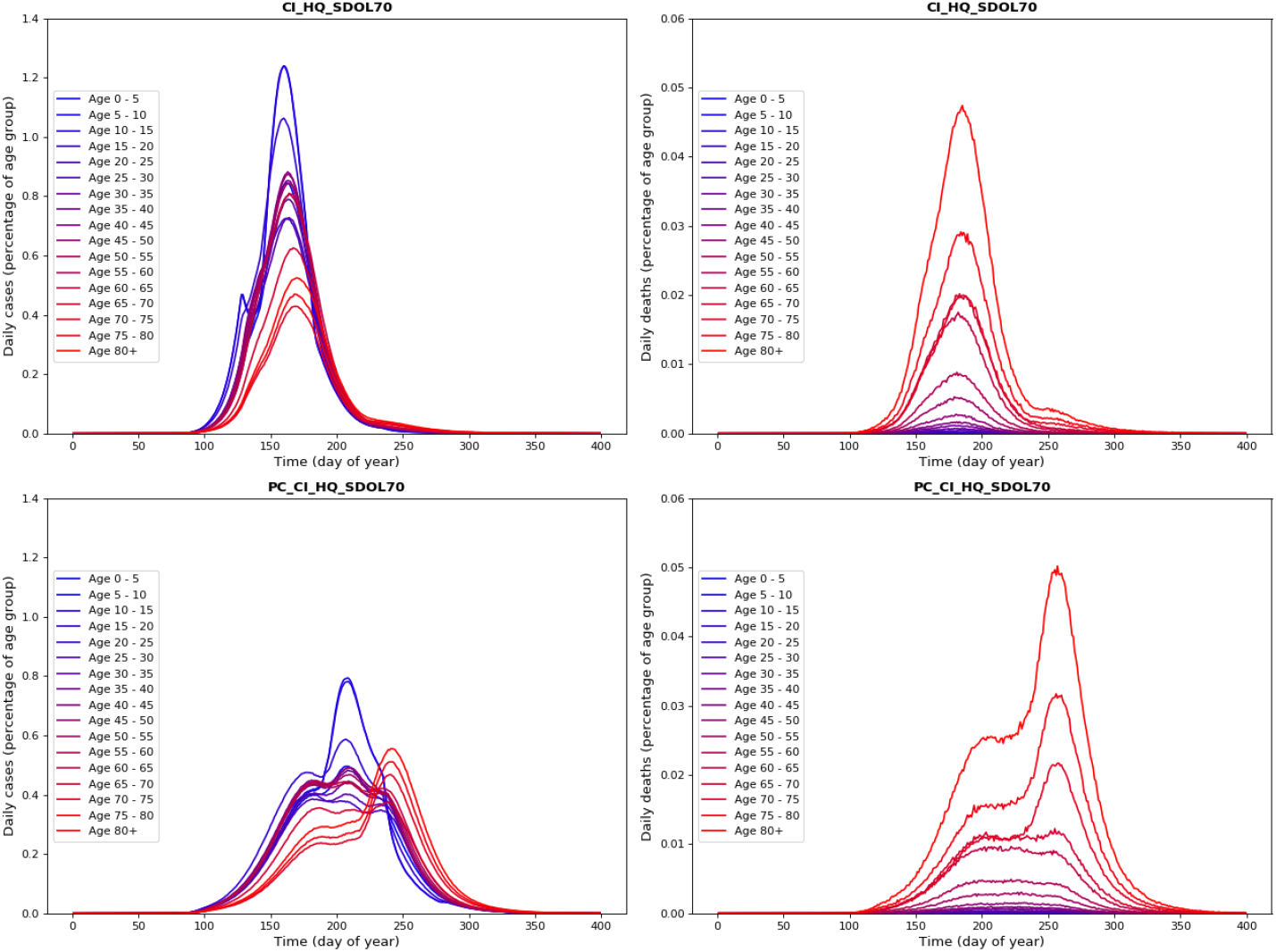
Simulated values for daily virus cases (left) and deaths (right), for scenarios CI_HQ_SDOL70 (top) and PC_CI_HQ_SDOL70 (bottom). Interventions are triggered by reaching 100 cumulative ICU cases. Results are broken down into age categories as indicated, with SDOL70 interventions affecting the three oldest groups. In the CI_HQ_SDOL70 scenario we see a single peak of cases, with greatest infection in the younger age groups but most deaths occurring in the older. In the PC_CI_HQ_SDOL70 scenario we see three peaks in the plot of daily cases, with the first peak occurring at a similar time for CI_HQ_SDOL70 above, but with reduced severity. The second peak seems to be a response to the ending of Place Closure (PC), and most affects the younger age groups, therefore having little impact on the total deaths. The third peak affects the older groups, leading to a significant increase in the total deaths. After the trigger, all the interventions are in place for 91 days the general social distancing runs to day 194, and the enhanced social distancing for over 70s runs for an extra 30 days.

### 4.1 CovidSim’s description of a second wave

Although Report 9 does discuss the possibility that relaxing the interventions could lead to a second peak later in the year, we wanted to briefly explore this in a bit more detail. Given that little data for intialising the model was available when Report 9 was released, we use an updated set of parameter files included in the github repository (8).

The interventions we consider are place closures (PC), case isolation (CI), household quarantine (HQ) and general social distancing (SD) which are implemented using the PC_CI_HQ_SD parameter file. These interventions start in late March (day 83) and last for 3 months (91 days). These simulations are also initialised so that there are about 10000 deaths by day 100 in all scenarios, presumed to have mostly been infected before the interventions were implemented.

The results are presented in Figure 4. The top panel shows cumulative deaths, with data from (11), while the bottom panel shows ICU bed demand per 100000 people. We consider a range of *R*_0_ values and find that values higher than those considered in Report 9 best reproduce the data, with *R*_0_ = 3.5 providing the best fit. This is consistent with the analysis presented in (7), but we acknowledge that the data could also be fitted by changes to the other scenario parameters. In both panels we also show the “Do nothing” scenario for *R*_0_ = 3.0.

**Figure 4:**
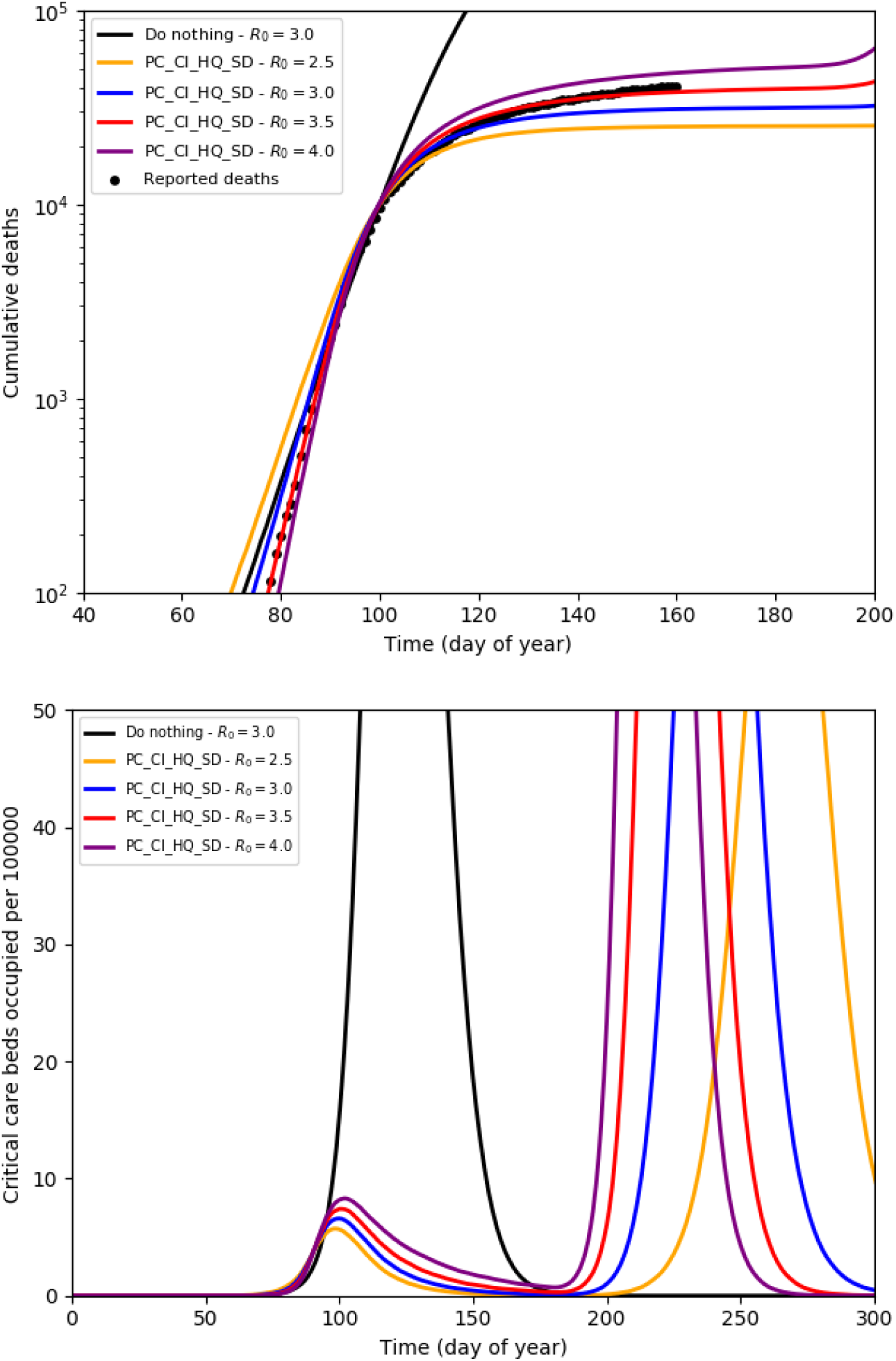
Refit of the CovidSim March parameterization based on death data through to June. The top panel shows cumulative deaths, with data from (11), while the bottom panel shows ICU bed demand per 100000 people. We considered a range of *R*_0_ values and find that values higher than that considered in Report 9 best reproduce the data. A good fit also requires us to assume that the epidemic started considerably earlier than was previously suggested in Report 9. We see that CovidSim (a) provides a good fit to the data with a value of *R*_0_ = 3.5 and (b) correctly predicts that the ICU capacity would not be exceeded during the intervention period.

The ICU bed demand for the scenarios presented in Figure 4 show that the interventions are correctly predicted to limit ICU demand to less than 10 per 100000. However, if interventions are fully lifted, there is a risk that this demand will start increasing again sometime between day 200 and day 250 (i.e., between late July and late August). If some interventions were to remain in place, then this might delay, and weaken, this second wave.

As illustrated here, and in our analysis of the mitigation scenarios in Report 9, it would seem important to understand how the intervention scenario, and the subsequent relaxation of the imposed interventions, may influence a potentially more deadly second wave that could occur later this year.

## 5 Conclusion

In this paper we used the recently released CovidSim code (8) to reproduce the mitigation scenarios presented in mid-March in Report 9 (4). The motivation behind this was that some of the results presented in Report 9 suggested that the addition of extra interventions may actually increase the total number of deaths.

We find that the CovidSim code reliably reproduces the results from Report-9, and that the model underlying CovidSim can accurately track the UK death-rate data. To do so does require an adjustment to the parameters, a slightly higher *R*_0_ than considered in Report 9, and results in an earlier start to the epidemic than suggested by Report 9. We emphasize, though, that the unavailability of these parameters in early-March is not a failure of the model.

We confirm that adding school and university closures to case isolation, household quarantine, and social distancing of those over 70 would lead to more deaths when compared to the equivalent scenario without school and university closures. Similarly, adding general social distancing to a case isolation and household quarantine scenario was also projected to increase the total number of deaths.

The qualitative explanation for this is that within all mitigation scenarios in the model, the epidemic ends with herd immunity with a large fraction of the population infected. Strategies which minimise deaths involve having the infected fraction primarily in the low-risk younger age groups. These strategies are different from those aimed at reducing the ICU burden.

A key thing seems to be that scenarios that are very effective when the interventions are in place, can then lead to a second wave during which most of the infections, and deaths, occur. Our comparison of updated model results with the published death data suggests that a similar second wave could occur later this year if interventions are fully lifted.

Finally, we reemphasize that the results in this work are not intended to be detailed predictions for the second wave. Rather, we are reexamining the evidence available from CovidSim at the start of the epidemic. More accurate information is now available about the compliance with lockdown rules and age-dependent mortality. The difficulty in shielding care-home residents is a particularly important piece of health data that was not available to modellers at the outset. Nevertheless, in almost all mitigation scenarios, CovidSim epidemics eventually finish with widespread immunity, and the final death toll depends primarily on the age distribution, not the total number, of infections.

## Data Availability

Code data is publicly available at https://github.com/mrc-ide/covid-sim/

## Acknowledgements

We acknowledge support from UKRI grant ST/V00221X/1 under COVID-19 initiative. This work was undertaken [in part] as a contribution to the Rapid Assistance in Modelling the Pandemic (RAMP) initiative, coordinated by the Royal Society.

## Supplemental Information

**Figure 5:**
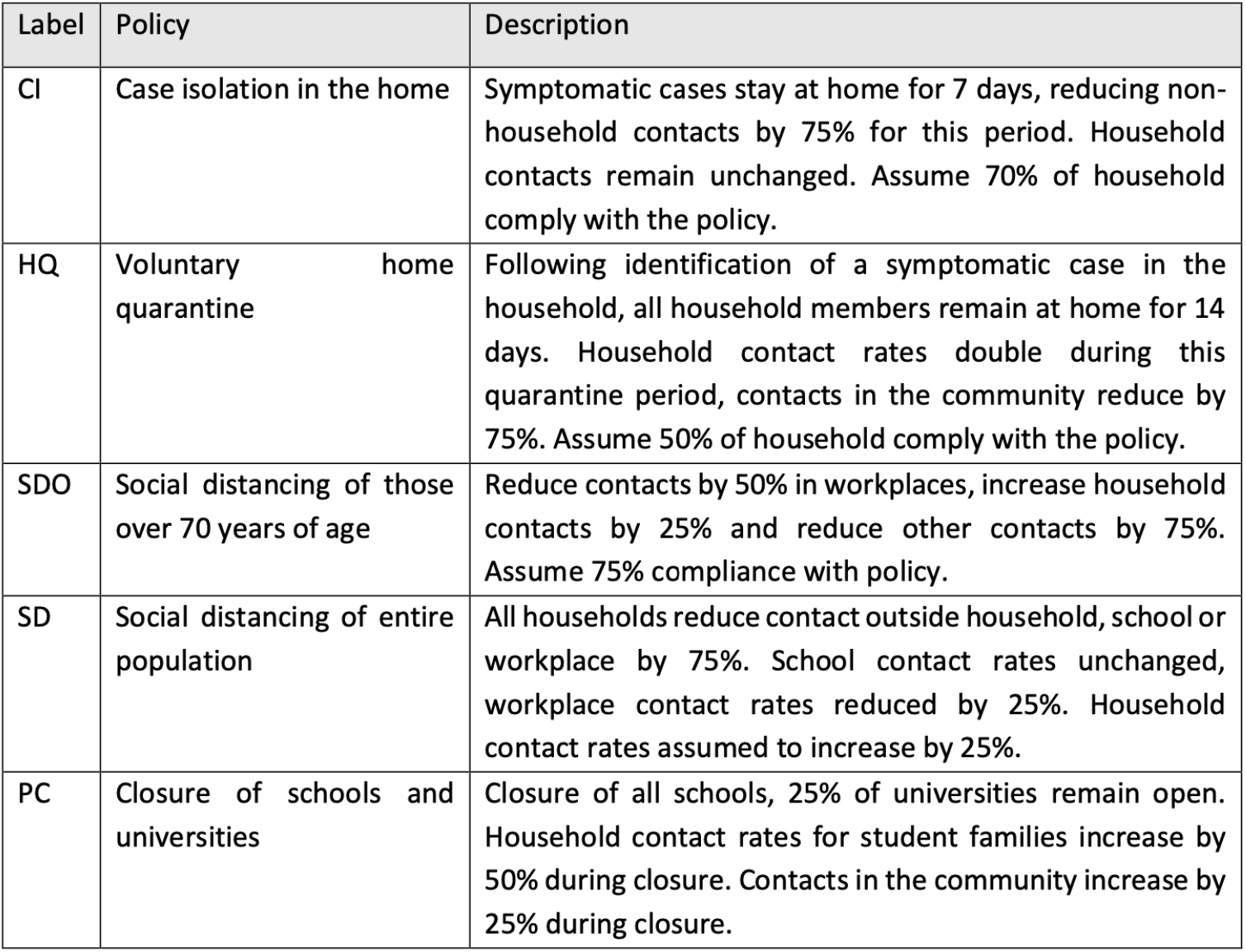
Table defining the interventions considered in CovidSim copied from Report 9.

## Footnotes

1 throughout this paper, we maintain the distinction between epidemiological “model”, and software implementations as “code”

2 Despite the description of place closure interventions in Table 2 of Report 9, university closures are not included in the (PC_)CI_HQ_SDOL70 scenario parameter files (8)

